# Use of Modeling to Inform Decision-Making During the COVID-19 Pandemic: A Qualitative Study

**DOI:** 10.1101/2021.04.13.21255401

**Authors:** Karl Johnson, Caitlin Biddell, Kristen Hassmiller Lich, Julie Swann, Paul Delamater, Maria Mayorga, Julie Ivy, Raymond Smith, Mehul Patel

## Abstract

**Background:** The COVID-19 Pandemic has popularized computer-based decision-support models as a tool for decision-makers to manage their organizations. It is unclear how decision-makers have considered these models to inform COVID-19-related decisions.

**Methods:** We interviewed decision-makers from North Carolina across diverse organizational backgrounds to assess major decision-making processes during COVID-19, including the use of modeling as an input to inform decision-making.

**Results:** Interviewees were aware of models during COVID-19, with some depending upon multiple models. Models were used to compare trends in disease spread across localities, allocate scarce resources, and track disease spread within small geographic areas. Decision-makers desired models to project disease spread within subpopulations and estimate where local outbreaks could occur as well as estimate the outcomes of social distancing policies, including consequences beyond typical health-related outcomes. Challenges to the use of modeling included doubts that models could reflect nuances of human behavior, concerns about the quality of data used in models, and the limited amount of modeling at the local level.

**Conclusions:** Throughout COVID-19, decision-makers perceived modeling as valuable for understanding disease spread within their communities and to inform organization decisions, yet there were variations in organizations’ ability and willingness to use models for these purposes.

## Introduction

The COVID-19 Pandemic has challenged decision-makers to manage their organizations through a public health crisis.^1–3^ Many organizations have shifted the type of services offered, establish structures for emergency decision-making, and develop the capacity to track and act upon continuously updated COVID-19-related information.^4,5^ In parallel, the pandemic has accelerated health sciences research and popularized many tools of health science researchers,^6–9^ including computer-based decision-support models designed to simulate the future course of disease spread under different epidemiologic and policy assumptions.^10,11^ Throughout the pandemic, it has been unclear how decision-makers across a range of organizational backgrounds (especially those outside Public Health or Healthcare) have used these models. In this qualitative study, we aimed to better understand the use of COVID-19 models for organization-level decision-making.

## Methods

From October 2020 to January 2021, we interviewed decision-makers (n = 44) from North Carolina across diverse organizational backgrounds, including three or more interviewees from each of the following eight sectors: education (primary/secondary and higher), public health, public safety, government, healthcare, business, faith-based, and public transportation. Through semi-structured interviews, we assessed their organization’s major decision-making processes throughout the pandemic. A codebook was developed using an inductive approach to label responses across several broad categories, including decision inputs for COVID-19-related decisions. All interviews were double coded independently by two members of the research team. In this analysis, we focused on coded segments discussing the use of modeling as an input to inform decision-making, and identify key themes to help modelers appreciate current perceptions about modeling and opportunities to improve the use of modeling in decision-making. As asked in the interview, different types of simulation-based models were not distinguished (e.g. curve-fitting models, agent-based simulations, compartmental models, etc.), but were treated as a general category.

This study was exempted from IRB review by the *[blinded-for-review]* Office of Human Research Ethics.

## Results

Among the 44 decision-makers interviewed, twenty substantively discussed modeling as a decision input (**Table 1**); others were either not aware of modeling or had not considered it as an input to decision-making. Comments among this subset were categorized into instances of actual use, recommended applications, and resistance toward model-informed decision-making during COVID-19. Representative quotes across themes are in **Table 2**.

**Table 1:** Distribution of Interviewees by Sector (N=20*)

| **Table 1: Distribution of Interviewees by Sector (N=20\*)** |  |  |
| --- | --- | --- |
| **Sector** | **Number of Interviewees** | **Organization roles included** |
| Public Safety | 2 | County Sheriff, Director of County Emergency Services |
| Healthcare | 2 | Systems Engineer for Private Health System, President of Healthcare Association |
| Public Health | 4 | Director of Local Health Department |
| Education | 7 | Director of University Emergency Management, County School Board Member, Elementary School Principal, Director of University Student Health Services, Superintendent of County Public School System, Vice Provost for University Enrollment Management, Community College President |
| Faith-based | 3 | Catholic Priest, COVID-19 Task Force Lead at a Catholic Parish, Presbyterian Minister |
| Government | 2 | County Manager, Assistant County Manager |

*\*This number reflects the amount of decision-maker interviewees with substantive commentary on modeling, either with respect to their own use of modeling, desired uses for modeling, or points of resistance to model-informed decision-making.*

**Table 2:** Major Interview Themes and Representative Quotes.

| *Major Theme* | *Sub-theme* | *Representative quote(s)* |
| --- | --- | --- |
| Use of Modeling during COVID-19 | Use of multiple models to inform decision-making | “...we probably had about five different models that covered not only our state, and our local area and also compared that to the national level...We weren't going to just isolate or pick and choose certain things.” (Healthcare, E3)<br>“We have three different model sets that we would watch. One is our own internal modeling system...We would constantly compare their models to ours and where we were actually...We used those [national models] to...make sure there was some type of sanity check with the protective actions that we were putting in place here locally.” (Public Safety, H3) |
|  | Importance of modeling surge capacity | “The hospital surge capacity is where we start to get concerned. Any modeling that we see that shows that the hospital capacity is decreasing then we start to absolutely be concerned.” (Public Safety, H5) |
|  | Using modeling to compare trends between localities | “[Modeling] allowed us to compare ourselves to other communities of similar size and density...[it] gave us a real gut check on how we were doing compared to other communities versus just the rest of North Carolina.” (Public Safety, H3)<br>“We were typically seeing ourselves about two weeks behind New York City, the West coast, Seattle, the major cities that were really blowing up.” (Healthcare, E3) |
|  | Using modeling to track the spread of disease within small geographic areas | “The modeling played a key role...with the different tools that we have from being able to graph who's getting infected to being able to show them by zip code what their infection rates look like.” (Public Health, D1) |
|  | Emphasis on the value of model visualizations | “We all learn from...visual cues. [Modeling is] not abstract. I mean, it is abstract, but it's not abstract words, it's that hard stuff you can hang your hat on...That was probably the most valuable thing...it's that visualization and just putting it in front of people's faces. Because otherwise, it's all words.” (School, C3)<br>“‘Here's what happens when you open it up.’ All of a sudden we see [the curve] go up. ‘Here's what you need to do if it gets too out of control.’...I appreciated it because even though it was the first time I seen it, it made sense. I was like, ‘Okay, we can see.’ For me, the goal for me was like, ‘Okay, how do I get it to where I don't have to expand and contract? I can slowly open it up.’ You get to the point where the curve just kind of like flattens out, and it even eventually starts going down.” (Public Health, D1) |
|  | Dependence on local health department for model assurance and results | “We weren't out trying to vet the data or peer review it or any of those kinds of things. But our health director was taking the data she received from the CDC, she was taking the information she received from the North Carolina Department of Health and Human Services, she was taking the models that they were using to create the guides that |
|  |  | they were giving. We took them to be trusted sources.” (Government, A3)<br>“No, we have not [used simulation models for decision-making], unfortunately not to my knowledge. I'd say we've used information from the local health department, and again, looking at infection rates and things of that nature.” (School, C4) |
| Recommended Applications for COVID-19 Modeling | Desire for models to predict disease spread within subpopulations | “We saw it go from a black African-American community to a Latinx and Hispanic community. And now it's gone to a white community...The modeling would be really interesting to see what was happening historically.” (Public Safety, H5)<br>“If we could know, like what cluster we are looking at next, what group of people could we potentially be looking at next. We can start tailoring our message and get out to those trusted leaders to say, "Look, you are not immune from this. This could very well affect you too." (Public Safety, H5). |
|  | Desire for modeling to inform safe re-opening strategies | “I would love to see the simulations, and this is what I think everybody would love to see, what happens if we get to a space where we fully reopen and we take the limits off of some of the occupancy/capacity...Would it be enough to do the masking and the hand-washing?...so maybe with the simulation and modeling, it would take into account if you had immunity amongst this percentage of the population because to me, maybe that would give us some goals in our vaccination planning and programs. ‘Well, if we get to this, we stand a better chance of this outcome.’” (Public Health D4).<br>“I think it'd be really useful. I'm a little worried we're not relying on them enough at this point in terms of designing our testing, procedures or plan, isolating plans, quarantining plans as we bring students back. Honestly, they may be as important in the ... important as we ramp up, but they're really important that when we hit a difficult area, that we don't make the wrong decision there. Can we work our way out of this problem versus, hey, send the kids home. How many kids do we actually have infected? How's it growing? Where is it?... I think we need more modeling to think about our testing plans. It is a challenge.” (School, C1) |
| Challenges with and Hesitancies toward COVID-19 Modeling | Hesitancy toward models given how sensitive they can be to model assumptions, leading to inconsistent results | “I've worked with models for a long time...And unfortunately, I know enough about models to make it dangerous for me and that knowing that if I change one little variable a little bit, I can predict the end of the world or the beginning of life.” (School, C3)<br>“It's just finding out or trying to put any signs we're using or modeling we're using in the proper context, knowing its |
|  |  | limitations and so forth because everyone's using it as like here's the answer. No, that's a possible answer certainly, but it may not be the answer, and what are the other possibilities of how accurate are these models. What's your point of failure? In what ranges of circumstances are they useful and outside of those ranges do they lose that? I'm still a bit puzzled on the whole thing." (School, C1)<br>"People want to see something that they can hang on. They can say, okay, this is what the model says. This is what's going to be accurate. And some of the models were so far apart that it was really challenging to come up with something specifically for [our county]." (Public Health D5)<br>I think as I mentioned earlier, one of the biggest challenges has been lack of consistency. Whether that's guidance, whether that's data use, whether that's modeling, whatever it happens to be. And as we go into discussions around vaccine, that consistent messaging is going to be huge or disastrous depending on which way it goes. So I think modeling is ... The work that they're doing is critical I think as we move forward on this because it's not going away anytime soon. And it would be incredibly helpful, but it would need to be consistent. And we need something that's clear, easily understandable by the general public. (Public Health D5) |
|  | Hesitancy that models can effectively incorporate the nuances of human behavior | "So those models could be helpful. But how do you factor behavior in those models? Assuming you can, and I do think you can, I mean, I do work with some of these models as well, and you can account for that kind of by different parameters in the model of that estimate how close people are in contact with each other, how many people are wearing masks, stuff like that. Compliance is the issue, I think." (Public Health, D5)<br>"So those models could be helpful. But how do you factor behavior in those models? Assuming you can, and I do think you can, I mean, I do work with some of these models as well, and you can account for that kind of by different parameters in the model of that estimate how close people are in contact with each other, how many people are wearing masks, stuff like that. Compliance is the issue, I think." (School, C5)<br>"...it would be helpful to understand the human behavior, reality of truth, how its impacted the success of slowing the curve or flattening. Because I think that a lot of the conversations that we have now are complacency driven." (School, C2) |
|  | Hesitancy about the data used to develop models | "If we had good data, modeling would be great. It would help us look over time at what's happened here...to predict what could potentially happen moving forward based on |
|  |  | changes that we're seeing and what's reopening, how many ... What percentage of people are actually wearing masks. There are all sorts of things that could feed into that modeling if we had good data. But it's a little challenging right now." (Public Health, D5)<br>"I think modeling would help... I think we need to be a lot more transparent with the data to comfort the people." (School, C1) |
|  | Inability of state-level models to translate to individual counties | "The other problem was a lot of the models are at the very best are state level models. They're not local. And because we looked pretty different than the rest of the state in terms of numbers and those sorts, they wanted to see something from here. So it made it challenging with the models." (Public Health, D5)<br>"Well, and I will tell you that it's hard for us. We look at the state data, but we look more at our local data just because we don't look like the rest of the state...The modeling for me would be very helpful if there was a focus on what's going on here. It's important for me to know what's going on in the state. Don't get me wrong there, but at the same time, it doesn't help me to look at the state and see that their numbers are going one way and mine are going the other." (Public Health, D5) |

### Use of Modeling during COVID-19

Decision-makers consistently demonstrated awareness of COVID-19 decision-support models, with several following one or more local, state, or national-level models. Model sources varied among decision-makers. Whereas some organizations built their own internal models, most relied on local, external modeling groups. In the absence of local modeling resources, several decision-makers relied on national models, especially those from the Institute for Health Metrics and Evaluation (University of Washington), to inform initial decision-making around shut-downs and safety protocols. One decision-maker from the healthcare sector commented on the use of multiple models, with recognition that one model could not provide all the answers (Healthcare, E3). In other instances, decision-makers used external models for comparison with internal models, so as to check that decision-making at the local level made sense in the context of state-or national-level trends (Public Safety, H3). This aligns with a general wariness, as expressed across most interviewees, toward making a decision that would draw negative attention to their locality or organization if their local model was wrong.

Three healthcare organizations utilized models for allocating scarce resources and critical care employees. Among healthcare system decision-makers and others, hospital surge capacity models were the most commonly used models for decision-making (Public Safety, H5). In two other instances, decision makers used modeling to monitor disease spread in comparable localities to estimate what could be expected in their own jurisdiction (Public Safety, H3). Comparisons occurred not just within North Carolina, but also across other states, especially those that had been hardest hit by the Pandemic early on (Healthcare, E3). Three other respondents emphasized how valuable modeling was for tracking the spread of disease within small geographic areas, especially those that correspond with vulnerable communities that could be reached through targeted communication, testing, and contact tracing efforts. A local health department director, for instance, reflected on the importance of modeling infection rates by zip code (Public Health, D1).

There were two instances in which decision-makers emphasized the value of model visualizations and how they helped clarify the impact of policies on disease spread for multiple audiences. Visualizations helped to make the abstract complexity of models concrete (Education, C3). They were also educational. One Public Health official described the process of working with a government epidemiologist as they presented graphically how different policies may shift the “curve” of total infections over time in a way that was both intuitive and easily actionable (Public Health, D1).

Having someone in the organization who is trained to interpret models, point to key takeaways, and situate findings within the broader literature was a facilitator to using models to inform decision-making. In the absence of this internal knowledge, interviewees noted using models that had been approved by their local health director as a way to discern which models could be trusted (Government, A3).

### Recommended Applications for COVID-19 Modeling

There were multiple decision-makers who, while they had not used modeling at the time of the interview, proposed applications for which modeling would be useful. For instance, two individuals emphasized a desire for models to show disease spread within subpopulations, including a Public Safety officer who desired models to both understand how subpopulations with a high burden of disease shifted throughout the pandemic and to identify potential outbreaks (Public Safety, H5). Additionally, a public health officer wanted modeling to help show the outcomes of social distancing policies, and specifically to determine what minimum number of restrictions need to be in place to ensure a safe re-opening. They also proposed modeling to inform communication campaigns for vaccination rollout, with particular interest in identifying target coverage levels (Public Health, D4). A university administration officer similarly desired modeling to inform testing policies if schools were to return in-person (Education, C1). Another interviewee expressed a desire for modeling the downstream consequences of the pandemic beyond typical health-related outcomes, particularly assessing developmental outcomes for kids who have been in school virtually (Education, C7). In general, decision-makers used visual language (e.g. “show”, “see”) when describing their desire for future modeling work, and avoiding more technical words like “predict” and “forecast.”

### Resistance toward COVID-19 Modeling

Interviewees expressed resistance to modeling, including the recognition that models can be highly uncertain and sensitive to differing assumptions, which may lead to inconsistent results across different models (Education, C3; Education, C1; Public Health, D5). Points of resistance also included doubts that models could effectively incorporate the nuances of human behavior, particularly compliance with official mask usage guidelines (Public Health, D5; Education, C2; Education C5). Among such interviewees, there was an implicit assumption that models must rigidly mirror official guidelines (e.g. everyone wearing masks), without an appreciation for the flexibility models provide. Other decision-makers questioned the quality of data used in the models, acknowledging that “models are only as good as the comfort level you have with the data going into it and the assumptions being made” (Education, C3). This critique nevertheless acknowledged that models could become an effective tool for decision-making if quality data were used and transparently presented (Public Health, D5; Education, C1). Another concern raised was that models did not capture the nuances of local communities; decision-makers identified unique characteristics of their own communities which made them substantively different from the state at large. One local health director commented on the inability of state-level models to translate to individual counties (Public Health, D5).

## Discussion

Modeling has become a well-recognized tool for COVID-19-related decision-making within private and public organizations, including local organizations. Among decision-makers interviewed for our study, modeling was seen as valuable for understanding the trajectory of the pandemic within their communities and to inform organization decisions, such as opening facilities or allocating resources. Visualizations were valuable for understanding model outcomes. Notable points of resistance toward the implementation of modeling results included the difficulty of creating models which can be useful to local decision-makers accounting for differences across areas. Accurate modeling at the local level and modeling the impact of policy among subpopulations (especially vulnerable communities) both demand access to granular data which was difficult to obtain during COVID-19. ^12^ Examples include mobility data or data on mask usage, neither of which are easily accessible by age and race or ethnicity.^13^ Our study results emphasize the need to modernize the data infrastructure in the United States to support these model development and calibration needs. In the absence of internal modeling capacity and data collection locally, local decision-makers may have to rely on externally built models that may not take into account important features of the local context.^14^ Given the resistances raised, additional guidance is needed to help local decision-makers become better consumers of modeling, including the capacity to understand decision objectives, potential model approaches and outcomes (e.g., predictions, forecasts), the uncertainty associated with model structure and estimates, and effective communication of model-informed decisions.^15^

## Data Availability

Qualitative Data used for this analyses may be accessed upon request.

